# Prevalence, Genetic Diversity, and Landscape Associations of *Orthohantavirus puumalaense* in Bank Voles (*Clethrionomys glareolus*) from Northern Sweden

**DOI:** 10.64898/2026.06.02.26354689

**Authors:** Alina Johanna Anton, Rainer G. Ulrich, Valerie Allendorf, Hannes Bergmann, Lutz Breuer, Zihan Dai, Stephan Drewes, Arne Hegemann, Yonas Meheretu, Frauke Ecke, Sascha Knauf

## Abstract

Puumala hantavirus (PUUV, *Orthohantavirus puumalaense*) is one of the primary causative agents of haemorrhagic fever with renal syndrome in Europe and is maintained in natural populations of the bank vole (*Clethrionomys glareolus*, also known as *Myodes glareolus*). Despite public health relevance, we are only starting to understand the molecular properties and interplay between environmental and ecological factors of the pathogen that explain PUUV infection in bank voles. Here, we investigated PUUV occurrence, genetic structure, and environmental associations in bank voles sampled from two boreal forest areas in northern Sweden, during a complete vole population cycle (2020-2023). In total, 519 voles were screened for PUUV RNA using targeted reverse transcription PCR (RT-PCR). PUUV small (S-) segment RNA was detected in both study areas and observed infection patterns varied with sex, body weight, season and year. Specifically, we detected significant interactions between season and area and between season and body weight, with males showing consistently higher infection probabilities. Infection probability was also higher during periods of increased vole abundance and peaked in 2022. Phylogenetic analysis of partial S segment sequences demonstrated that all detected sequences clustered within the North-Scandinavian PUUV lineage, with no apparent spatial differentiation, indicating limited genetic structuring between the sampling areas. Habitat analyses at multiple spatial scales did not identify significant associations between PUUV occurrence and land-use variables, suggesting that infection dynamics were driven primarily by host demographic and temporal factors rather than broad-scale habitat composition. These findings highlight the importance of host demographics and temporal dynamics in shaping PUUV epidemiology in its reservoir, and provide additional insight into the molecular ecology of PUUV in northern Europe.

## 1 Introduction

The order *Bunyavirales* includes the family *Hantaviridae*, whose zoonotic members can cause haemorrhagic fever with renal syndrome (HFRS) and hantavirus pulmonary syndrome (HPS), also referred to as hantavirus cardiopulmonary syndrome (HCPS). Rodents are the main reservoir hosts for orthohantaviruses causing human infection, even though shrews, moles and bats were shown to represent reservoirs of additional hantaviruses with so far unknown zoonotic potential (Dafalla et al., 2022; Schlegel et al., 2012b). Although hantaviruses show broad evolutionary patterns consistent with long-term associations with their reservoir hosts, their diversification is not explained by strict co-speciation alone, host switching, local adaptation, and genome reassortment have been discussed as additional factors in hantavirus evolution (Plyusnin and Sironen, 2013).

Puumala hantavirus (PUUV, *Orthohantavirus puumalaense*) is a member of the family *Hantaviridae* and one of the principal causes of HFRS in Europe. This mild to moderate clinical form of disease caused by PUUV is also called nephropathia epidemica, (Lagerqvist et al., 2016). Like all orthohantaviruses, PUUV is an enveloped, negative-sense, single-stranded RNA virus with a tripartite genome comprising the large (L) segment (∼6.5 kilobases, kb), encoding the RNA-dependent RNA polymerase, the medium (M) segment (∼3.6 kb), encoding the glycoprotein precursor that is processed into Gn and Gc glycoproteins, and the small (S) segment (∼1.7 kb), encoding the nucleocapsid (N) protein (Bradfute et al., 2024). PUUV, as other cricetide-associated hantaviruses encode in an overlapping reading frame on the S segment an additional open reading frame (ORF) that encodes a putative non-structural (NSs) protein (Binder et al., 2021).

PUUV is primarily transmitted to humans through inhalation of aerosolised particles-contaminated excreta from infected bank voles (Vaheri et al., 2013) (*Clethrionomys glareolus*, formerly also known as *Myodes glareolus*) (Brummer-Korvenkontio et al., 1980; Vapalahti et al., 2003), the only confirmed reservoir host of PUUV. The bank vole is a common, generally forest-dwelling rodent widely distributed across Europe into central Asia (Wilson et al., 2017; Górska et al., 2025). The current distribution of PUUV and its genetic lineages is linked to the postglacial recolonization of Europe by bank voles from different refugia (Razzauti et al., 2009; Castel et al., 2019). Isolation of bank voles at these refugia likely resulted in the separation of different evolutionary lineages (Razzauti et al., 2009; Deffontaine et al., 2005). Central and Western Europe is thereby colonized by the Western, Carpathian and Eastern lineage of the bank vole (Drewes et al., 2017; Wójcik et al., 2010), whereas Sweden was colonized by two different routes: via the Ural lineage from the north and the Carpathian lineage from the south (Filipi et al., 2015). This recolonization resulted in the presence of the Central (CE) clade of PUUV in Western and parts of Central Europe, whereas bank voles in Fennoscandia are reservoirs of the Northern Scandinavian (N-SCA), Southern Scandinavian (S-SCA) and Finnish (FIN) lineage (Nemirov et al., 2010; Ling et al., 2024). In Europe, rodents and especially bank voles are known for their partly dramatic changes in population density. Bank vole populations in Fennoscandia and central Europe, undergo multiannual density fluctuations, often with 3-5-year cycles characterised by regularly recurring population peaks and declines (Hornfeldt, 1994; Lambin et al., 2025; Imholt et al., 2014; Tersago et al., 2008; Hornfeldt, 1994; Krebs and Myers, 1974; Hansson and Henttonen, 1985). These reservoir cycles are associated with temporal variation in number of notified nephropathia epidemica cases, since high bank vole abundance tends to result in high PUUV prevalence in bank voles and provides increased opportunities for virus transmission at the bank vole–human interface (Khalil et al., 2019; Reil et al., 2017; Wang et al., 2023; Kallio et al., 2009).

In Fennoscandia, PUUV transmission in bank voles is closely linked with bank vole demography, ecology, community structure, weather conditions, climate change, habitat structure and quality. Juvenile bank voles are protected from infection by maternal antibodies (Voutilainen et al., 2016; Kallio et al., 2006b). Higher PUUV prevalences in males than female (Voutilainen et al., 2012) can be attributed to males having larger homeranges than females (Bujalska, 1973; Mazurkiewicz, 1971).

An overall lower infection risk for bank voles can be associated with species-rich small mammal communities compared to those in species-poor communities (Khalil et al., 2016; Ecke et al., 2017). Habitat structure and quality affect occurrence and local persistence of PUUV (Magnusson et al., 2015; Khalil et al., 2017; Olsson et al., 2005). In addition, the environmental persistence of PUUV is affected by weather conditions (Kallio et al., 2006a), and pathogen transmission is higher during winters characterised by rainy instead of snowy early winters, which in addition to aggregated overwintering of bank voles, explains higher PUUV prevalence in spring than in autumn (Sipari et al., 2021). Although bank voles are typically chronically infected without overt clinical signs (Tscherne et al., 2025), overwintering survival and reproduction seem to be reduced(Kallio et al., 2007, 2015). In addition recent findings suggest immune modulation and a possible influence on the susceptibility for secondary infections (Schlohsarczyk et al., 2023). A fuller understanding of PUUV ecology in its natural reservoir requires direct molecular characterization of the virus itself, including its sequence diversity, phylogenetic structure, and potential associations with environmental factors.

In this study we therefore aimed to characterize the genetic diversity, phylogeny, and true prevalence of PUUV across two bank vole populations in boreal forest areas in northern Sweden. By integrating host demography, ecological data, and land-use patterns with viral genotyping, we sought to elucidate the factors determining viral distribution, prevalence and molecular evolution.

## 2 Material and Methods

### 2.1 Sampling Location and Samples

Bank voles from two sampling areas in Västerbotten County, northern Sweden, were included in this study. Both areas are located in a boreal forest landscape with the inland forest area near Vindeln (hereafter “Vindeln”, 64.2°N, 19.7°E) representing the middle boreal zone and the Coastal area near Umeå representing the transition between the middle and southern boreal zone (hereafter “Coastal area”, 63.8°N, 20.4°E) (Ahti et al., 1968).

All bank vole specimens in the Vindeln area were collected as part of the National Environmental Monitoring Programme of Small Mammals (NEMPSM) coordinated by FE and specimens are part of the National Environmental Specimen Bank (NESB) coordinated by the (The Swedish Museum of Natural History). The trapping at the Coast was neither part of the NEMPSM nor the NESB but followed the same approach. Bank voles were snap-trapped in permanent and systematically distributed 1-ha trapping plots (Vindeln: n = 58, Coast: n = 24). Each trapping plot consists of 10 trap stations, with five traps each, centred and spaced 10 m apart along the diagonal of the 1-ha square. Trappings were performed twice per year, each spring (May) and autumn (September), with each trapping session comprising three trap-nights. The trappings have been performed in Vindeln since 1971 and in the Coastal area since 2009 (Hörnfeldt, 1978; Hornfeldt, 1994). For this study, we used bank voles from spring 2020 to autumn 2023, comprising a complete vole cycle with 2020 representing the increase, 2021 the peak, 2022 the decline, and 2023 the low phase. During fieldwork, specimens were either kept in a mobile freezer (-20 °C) or in cool bags. Prior to permanent storage at -20 °C, species identification was performed and whole specimens were weighed. Individuals were then assigned to functional groups based on trapping season and body weight, with body weight thresholds being assigned after inspecting weight histograms: OWBR (overwintered breeders), defined as adults trapped in spring and weighing *≥*14.4 g, YBBR (year-born breeders), defined as non-adults trapped in spring and weighing < 14.4 g, SUBA (subadults), defined as individuals trapped in autumn and weighing < 21.5 g, and ADUL (adults), defined as reproductive or post-reproductive individuals trapped in autumn and weighing *≥* 21.5 g (Krebs and Myers, 1974; Ecke et al., 2002).

A total of 1,134 bank voles were dissected, including 823 individuals from Vindeln and 311 from Coast. The dissection and collection of organ material at the Swedish University of Agricultural Sciences followed a standardised protocol. Briefly, animals were thawed overnight at 4 °C prior to postmortem examination and dissected at room temperature. After sexing the animals, the right lung lobe was excised from the thoracic cavity, transferred into 2 mL CryoPure tubes (Sarstedt, Nümbrecht, Germany, cat.no.:72.379), and immediately stored at -20 °C . After 24 h the samples were then transferred to -80 °C freezer before shipping to the laboratories of the Friedrich-Loeffler-Institut (FLI). Samples were transported on ice and stored at ™80 °C at FLI before further processing.

The small mammal monitoring was approved by the Animal Ethics Committee in Umeå (Dnr A 18-2019), as well as by the Swedish Environmental Protection Agency (NV-07483-19), and all applicable institutional and national guidelines for the use of animals were followed.

### 2.2 Sample Size

To estimate PUUV RNA positivity in the two populations, a two-step screening strategy was applied. In this preliminary step, 40 individuals per area were tested. The apparent positivity observed during this initial screening was then used to calculate the required sample size for prevalence estimation in Epitools, assuming a test sensitivity and specificity of 0.99, an absolute precision of 0.05, and a confidence interval of 95% . Based on these calculations, at least 319 animals from Vindeln and 179 from the Coastal area were required to estimate true prevalence. To maximise extraction plate capacity, the final screened subset comprised 339 individuals from Vindeln and 180 individuals from the Coastal area. Induividuals selected for RT-PCR screening were drawn separatly for each area using simple random sampling without replacement in R v4.3.3 (Ooms, 2021) within RStudio v2026.01.0 (Posit team, 2025) using dplyr v1.1.4 (Wickham et al., 2023).

### 2.3 RNA Extraction

Lung samples were thawed on ice and a piece of 25 mg was cut and individually homogenised with a 5mm steal bead at 30 Hz for 180 s using a TissueLyser LT (QIAGEN, Hilden, Germany) following the manufacturer’s instructions for the NucleoMagVET Kit (Macherey-Nagel, Düren, Germany). Extraction was performed on a KingFisher Flex platform (Thermo Fisher Scientific, Waltham, MA, USA). Extracted RNA was transferred on ice from the extraction plate to 1.5 ml PCR clean SafeLock tubes (Eppendorf, Hamburg, Germany) and immediately frozen at -80 °C .

### 2.4 Mitochondrial DNA amplification

Bank vole lineage from ten animals per area was determined by sanger sequencing a fragment of the mitochondrial cytochrome b (cyt b) at Microsynth Seqlab (Göttingen, Germany). Briefly, conventional PCR amplified a 947 base pair (bp) fragment using universal primers as described by Schlegel et al. (2012a) in a total volume of 25 µL containing 2.5 µL template DNA using the GoTaq Flexi DNA Polymerase (Promega, Madison, WI, USA) according to the manufacturers instructions. The thermal cycling included an initial denaturation at 94 °C for 180 s, followed by 50 cycles at 94 °C for 30 s, 47 °C for 30 s, 72 °C for 60 s, and a final extension at 72 °C for 10 min. All PCR amplifications were run on a Biometra Trio thermal cycler (Analytik Jena, Jena, Germany). Amplicons were verified on 1.5% agarose gels for 50 min at 110 V, excised and purified using the QIAquick Gel Extraction Kit (QIAGEN, Hilden, Germany) according to the manufacturer’s instructions.

### 2.5 Hantavirus RT-PCR

A conventional S segment-specific RT-PCR targeting the nucleocapsid (N) protein-encoding region of PUUV and related hantaviruses was performed as described previously (Binder et al., 2020). Briefly, 2.5 µL of RNA was amplified in a final reaction volume of 25 µL using primers 342F (5^*′*^-TATGGTAATGTCCTTGATGT-3^*′*^) and 1102R (5^*′*^-GCCATDATDGTRTTYCTCAT-3^*′*^) and the SuperScript™ III RT/Platinum Taq Mix (Invitrogen, Karlsruhe, Germany). Reverse transcription was carried out at 50 °C for 45 min, followed by an initial denaturation at 94 °C for 120 s. Amplification was then performed over 40 cycles consisting of 94 °C for 30 s, 46.2 °C for 30 s, and 68 °C for 60 s, followed by a final extension at 68 °C for 10 min. RT-PCRs were run on a Biometra Trio thermal cycler (Analytik Jena, Jena, Germany). RT-PCR products were visualized on a 1.5% agarose gel. Amplicons of the expected size (760 nucleotides, nt) were excised and purified using the QIAquick Gel Extraction Kit (QIAGEN, Hilden, Germany) according to the manufacturer’s instructions. Amplification of beta-actin gene was included as an extraction control and was performed in parallel in the same PCR run (Wakeley et al., 2005).

### 2.6 Sanger sequencing and data analysis

Amplicons were Sanger sequenced at least once in each direction by Microsynth Seqlab (Göttingen, Germany). Sequence chromatograms were inspected manually, ends with an error probability >5%, as well as primer binding sites were trimmed. Consensus sequences were generated from forward and reverse reads in Geneious Prime v2025.1.3 (Biomatters Limited, Auckland, New Zealand) and compared against GenBank entries using blastn. Partial PUUV S-segment consensus sequences were aligned with the PUUV reference strain (PUUV strain Puu/Vindeln/L20Cg/83, Accession No. Z48586.1) and 126 unique hantavirus sequences retrieved from GenBank (Supplementary Table 1) using Multiple Sequence Comparison by Log-Expectation (MUSCLE) ((Edgar, 2004), version5.1) in Geneious Prime using the Parallel Perturbed Probcons (PPP) algorithm with 0 Hidden Markov Model (HMM) perturbations, without guide tree construction, and using 4 threads. Sequences covering a 685-nt region of the PUUV S segment were retained for phylogenetic analysis.

Bank vole cytochrome b sequences covering an 834-nt region were aligned with a reference sequence (Accession No. AF429810.1) and representative sequences from each lineage (Drewes et al., 2016).

A maximum-likelihood phylogeny was inferred using IQ-TREE v2.4.0 (Trifinopoulos et al., 2016) applying automated model selection (ModelFinder) (Kalyaanamoorthy et al., 2017), and branch support assessment using SH-aLRT with 1,000 replicates as well as ultrafast bootstrapping (1,000 replicates) (Hoang et al., 2018). The resulting tree was visualized using Interactive Tree of Life (iTOL) v7 software (Letunic and Bork, 2024) and lineage info, geographical origin and the amount of identical sequences collapsed into unique sequence types.

In addition, geographic distances were calculated from sampling coordinates using the Haversine formula, and their association with genetic distances was assessed with a Mantel test using vegan v2.6-4 (Oksanen et al., 2022) in RStudio.

### 2.7 Statistics

#### 2.7.1 Prevalence

RT-PCR outcome prevalence was calculated as the proportion of positive samples among all tested individuals, overall and by area, sex, body weight, season, and year. 95% confidence intervals were estimated using the Clopper–Pearson method. Individuals sampled in 2023 and weighing < 14.4 g were excluded from the general linear model (GLM) analysis because these groups were sparsely represented and contained no PUUV-positive cases. This resulted in sparse data structure and unstable model estimation, preventing reliable estimation of their effects. This threshold removed all YBBR individuals and a subset of SUBA individuals with body weights below 14.4 g, while OWBR and ADUL individuals were retained by definition. We modelled PUUV RNA occurrence (presence/absence) using a GLM with a binomial error distribution and logit link function. Predictors included Area (Vindeln vs. Coast), Season (Spring vs. Autumn), Sex (Male vs. Female), standardized body weight, and Year (2020–2022). The initial full model included all two-way interactions among predictors. Model reduction was performed from the full model using backward stepwise selection based on Akaike’s Information Criterion (AIC). The significance of predictors retained in the final model was assessed using likelihood-ratio tests (LRTs). Model-predicted RT-PCR positivity and 95% confidence intervals were calculated for each group using estimated marginal means (emmeans v1.11.2-8 (Lenth, 2025)) on the probability scale. Where the *Season* ×*Area* interaction was significant, within-area seasonal contrasts were computed with Holm adjustment, otherwise, an overall contrast was reported. Predicted prevalence was visualised with 95% confidence intervals using the ggplot2 v4.0.2 (Wickham, 2016). All statistical analysis were performed in R within RStudio.

#### 2.7.2 Land-use

Land cover data were obtained from the Swedish National Land Cover Database (NMD, NMD2023, version 0.1) that has a 10 m resolution and minimum mappable unit of 10 square meters. For each trapping plot, habitat composition was quantified within 2.5×2.5 *km* and 200×200 *m* polygons centred on the plots, and the proportional cover of each land-cover class was extracted. For similar approaches on the time series data, see (Ecke et al., 2017, 2006, 2013). We used two sets of NMD data: Dataset 1) we grouped the original 52 NMD classes (Supplementary Table 2) into broader habitat categories: Arable land (NMD code 3), artificial land (i.e., built-up or otherwise human-made surfaces and associated areas, NMD code 51-54), Water (NMD code 61-62), Coniferous forest (NMD code 111-113, 121-122) , Mixed forest (NMD code 114,123-124), Deciduous forest (NMD code 115-117,125-127), Clear-cuts (NMD code 118,128), Wetland (NMD code 200, 211-228), and Open land (411-413, 4211-4213, 4221-4223, 4231-4233). Dataset 2) we identified ecologically important individual NMD classes for bank vole and PUUV occurence and abundance among the original NMD classes, which included the NMD classes Clear-cuts, Coniferous forest, Mixed forest, Deciduous forest, Wetland as well as Buildings (NMD code 51) (Khalil et al., 2014; Huitu et al., 2003; Savola et al., 2013; Voutilainen et al., 2012; Olsson et al., 2005). The spatial analyses were performed in QGIS v3.40.5 (QGIS Association, 2026) were land cover per polygon was extracted with the function ‘Zonal histogram’.

PUUV occurrence (presence/absence) (Supplementary Table 3) was analysed in relation to habitat composition using individual-level generalised linear mixed-effects models (GLMMs) with a binomial error distribution and logit link. Models were fitted in R using the glmer function in the lme4 package v1.1-35.1 (Bates et al., 2015). PUUV infection status was used as the binary response variable, while season, habitat composition variables, and the sampling intensity per trapping plot were included as predictors. Trapping plot (TOPO) was included as a random intercept to account for non-independence of individuals originating from the same plot.

Proportional habitat variables were standardised prior to model fitting to improve numerical stability and allow comparison to effect sizes. Water was excluded to avoid perfect multicollinearity among the land cover data and because it was not considered suitable habitat for bank voles. The full model at the 2.5×2.5 *km* scale included Season, Artificial, Mixed Forest, Wetland, Coniferous Forest, Decidious Forest, Clear-Cut, Open Land, Arable Land and the sampling intensity per trapping plot. Multicollinearity was assessed using the variance inflation factors (VIF) from the car package (Fox and Weisberg, 2019), with a cutoff of <3.3.

At the 200×200 *m* scale, clear-cut caused severe multicollinearity in the coarse habitat model and was therefore excluded from the 200×200 *m* coarse-scale analysis. After exclusion, VIF values for the retained predictors were <2.

In the second step, we used NMD dataset 2 to build an expert-based model. This model included Season, Building (part of the Artificial predictor), Clear-Cut, Coniferous Forest, Deciduous Forest, Mixed Forest, Wetland and the sampling intensity per trapping plot. At the 200× 200 *m* scale, clear-cut also inflated VIF values in the expert-based model and was therefore excluded from the 200 ×200*m* expert-based analysis. Models were fitted using the bobya optimizer with increased iteration limits.

To provide ecological context for the habitat analyses, we conducted a explorative descriptive multiscale landscape assessment comparing PUUV-positive and PUUV-negative trapping plots with the broader landscape background. Random points were generated within a study area defined by a 50 km buffer around all trapping plots and clipped to the Swedish mainland. Land-cover composition was extracted from the original ungrouped NMD classes using circular buffers of 113 m, 564 m, and 1410 m radius (areas corresponding to 200 m, 1 and 2.5 km squares, respectively) centred on trapping and random points (Supplementary Figure 1), representing local habitat conditions, intermediate landscape context, and broader landscape structure relevant to bank vole ecology and PUUV transmission. Land-cover composition was compared descriptively among PUUV-positive, PUUV-negative, and random points using grouped boxplots (Supplementary Figure 2). The analysis was exploratory and not intended for inferential testing. They were implemented in R using sf, terra and tmap as key packages.

## 3 Results

### 3.1 Bank vole population dynamics and phylogeny

The small mammal monitoring in spring 2020 to autumn 2023 in Vindeln and the Coastal area yielded 2,720 bank voles (Vindeln: 1,818, Coast: 902). The population dynamics reveal that the population cycle started with an increase phase in spring 2020 that transitioned into a peak phase in 2021 and a decline phase in 2022, while spring 2023 initiated the low phase. The subset tested for PUUV RNA comprised of 133 females and 206 males in Vindeln (Figure 1), including 46 OWBR, 3 YBBR, 73 SUBA, and 11 ADUL females, and 87 OWBR, 111 SUBA, and 8 ADUL males. In the Coastal area subset, 78 females and 102 males were included, comprising 15 OWBR, 54 SUBA, and 9 ADUL females, and 42 OWBR, 51 SUBA, and 9 ADUL males. No YBBR individuals were present in the Coastal area subset.

**Figure 1.**
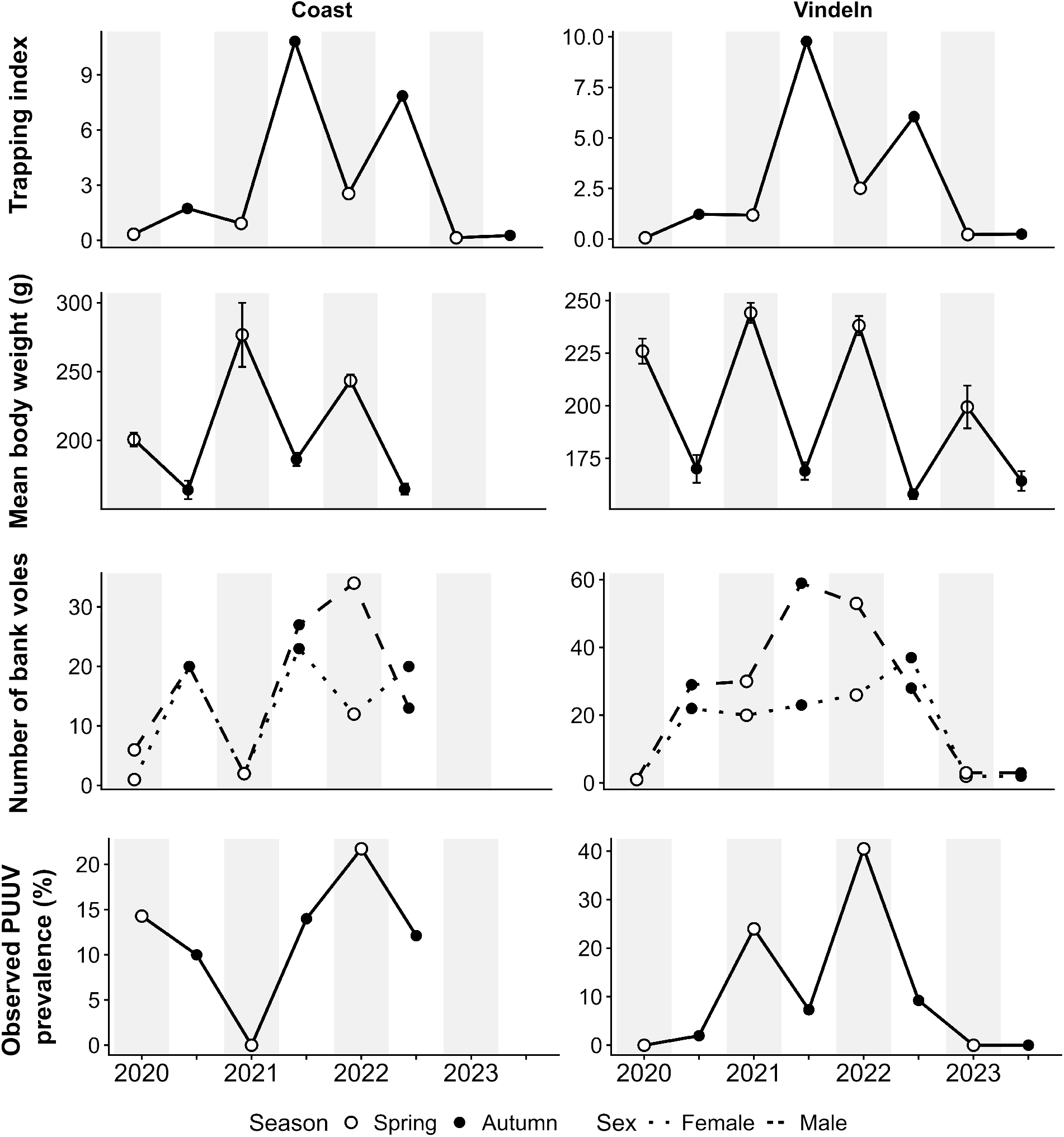
Temporal variation in trapping index, demographic composition, and observed PUUV prevalence in bank voles sampled from 2020 to 2023 in boreal forests of northern Sweden. Trapping index was calculated using all traps and captured animals in the sampling areas, while demographic dynamics and PUUV prevalence refer only to individuals included in the PUUV screening. No individuals from the coastal area were tested for PUUV in 2023. Spring values are shown as white circles and autumn values as filled black circles.

Cyt b sequence determination indicated that all bank voles belong to the Ural lineage.

### 3.2 Hantaviruses

The initial screening of 40 individuals per site suggested apparent PUUV RNA positivity of 27.5% (11/40) in Vindeln and 12.5% (5/40) in the Coastal area. Based on these result the sample size for the extended screening was calculated. Extended screening identified 83 PUUV RNA-positive individuals among 519 tested bank voles, comprising 57/339 in Vindeln and 26/180 in the Coastal area. Unadjusted observed prevalences, show seasonal fluctuations with highest observed prevalences in spring 2022, i.e. the decline phases in both sampling areas (Figure 1).

The final GLM retained Sex, Year, the *Season*×*Area* interaction, and the *Season*× *Bodyweight* interaction. PUUV positivity was higher in males than females and increased with body weight. A significant *Season* ×*Area* interaction indicated differing seasonal infection patterns between study areas, with the effect of body weight varying between seasons and the decline year 2022 showing the highest PUUV occurence proability (Table 1), 2).

**Table 1.** Likelihood-ratio tests for terms retained in the final GLM including bank voles >14.4g from Vindeln and coastal area from the years 2020-2022.

| Coefficient | Df | Deviance | AIC | LRT | Pr(>Chi) |
| --- | --- | --- | --- | --- | --- |
| <none> |  | 360.93 | 378.93 |  |  |
| Year | 2 | 71.30 | 385,30 | 10,37 | **0.006** |
| Sex | 1 | 377,42 | 393,42 | 16,48 | **<0.001** |
| Season x Area | 1 | 369,88 | 385,88 | 8,95 | **0.003** |
| Season x Bodyweight | 1 | 371.04 | 387.04 | 10.10 | **0.002** |

Bank voles sampled in 2023 and individuals <14.4g were excluded from the GLM, the dataset used comprised 285 individuals >14.4g from Vindeln, of which 57 tested positive (females: 13/107, males: 44/178), and 158 individuals >14.4g from Coast, of which 26 tested positive (females: 4/66, males: 22/92).

**Table 2.** GLM for PUUV occurence with five predictors including bank voles >14.4g from Vindeln and coastal area from the years 2020-2022.

| Coefficient | Estimate | Std.Error | z value | Pr(> \|z\|) |
| --- | --- | --- | --- | --- |
| Intercept | -2,68 | 0,53 | -5,07 | **<0.001** |
| Season (Spring vs. Autumn) | -0,78 | 0,55 | -1,41 | 0,157 |
| Area (Vindeln vs. Coast) | -0,67 | 0,42 | -1,60 | 0,109 |
| Year (2021 vs. 2020) | 0,44 | 0,51 | 0,87 | 0,385 |
| Year (2022 vs. 2020) | 1,30 | 0,52 | 2,47 | **0.014** |
| Sex (Male vs. Female) | 1,23 | 0,32 | 3,82 | **<0.001** |
| Body weight | 1,05 | 0,27 | 3,90 | **<0.001** |
| Season(Spring) x Area(Vindeln) | 1,74 | 0,59 | 2,95 | **0.003** |
| Season(Spring) x Bodyweight | -1,18 | 0,37 | -3,17 | **0.001** |

Reference levels were autumn, coast, 2020, and female. Body weight was z-standardized, therefore, the intercept represents the predicted log-odds of PUUV positivity at mean body weight. Main effects involved in significant interactions are conditional on the reference levels.

Estimated marginal means from *Season*× *Area* of the final model showed adjusted predicted prevalences in the Coastal area of 9.4% (95% *CI* : 4.0 −20.5) in spring and of 18.4% (95%*CI* : 10.9 −29.3) in autumn, whereas in Vindeln it was 23.0% (95% *CI* : 13.2 −37.2) in spring and 10.3% (95% *CI* : 5.8− 17.7) in autumn. Within-area contrasts indicated no significant seasonal difference in the Coastal area, but a significant seasonal difference in Vindeln, where the odds of PUUV RNA positivity were lower in autumn, than in spring (*OR* = 0.38, 95% *CI* : 0.16 − 0.92, *z* = 2.16, *p* < 0.05) (Figure 2 (a)).

**Figure 2:**
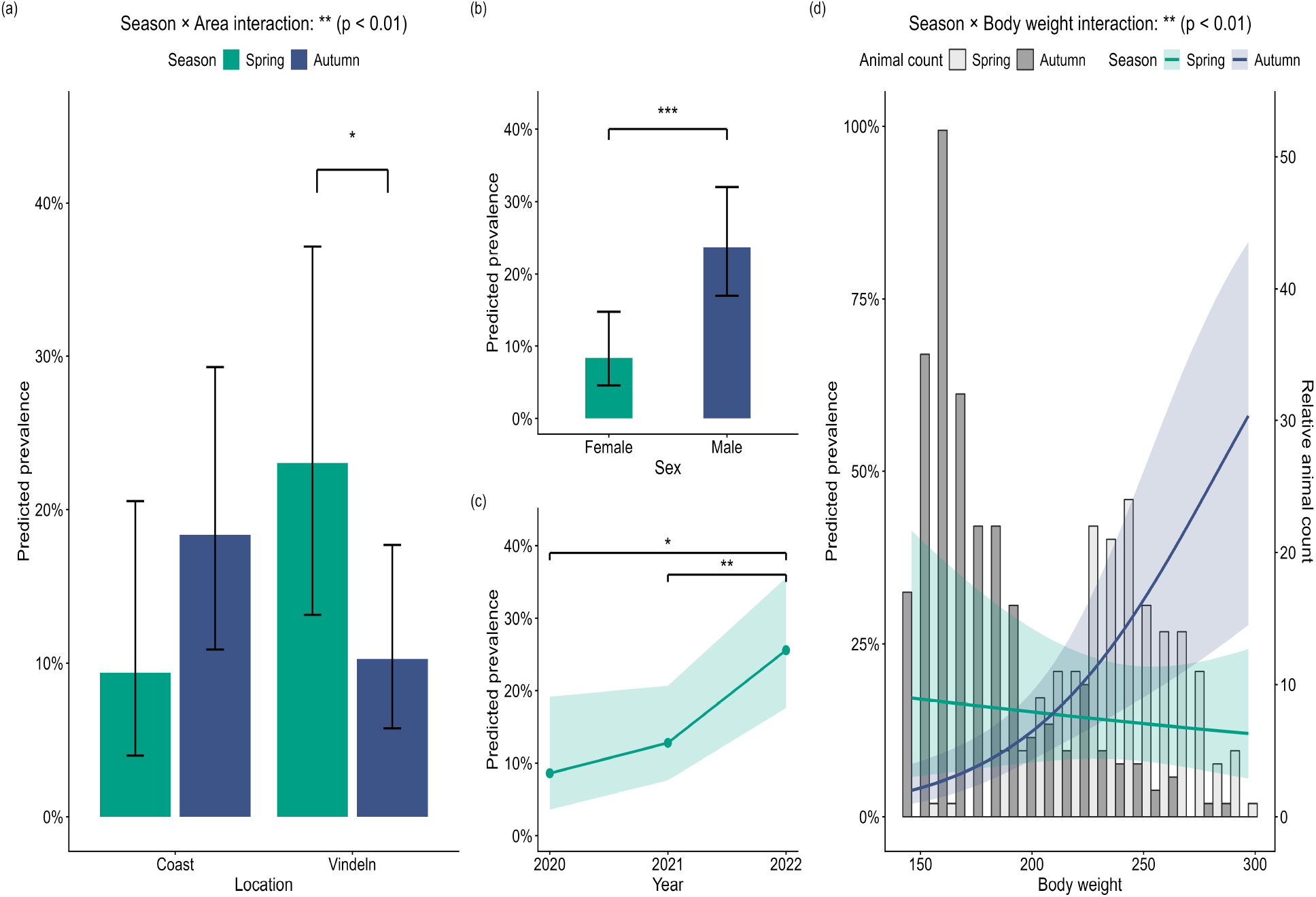
Predicted PUUV prevalence from the final binomial GLM including bank voles >14.4g from Vindeln and coastal area from the years 2020-2022: Panel (a) shows predicted prevalence by Season and Area, panel (b) by Sex, panel (c) by Year and panel (d) by season and Body weight including trapping counts in light grey. Bars, points, and lines represent model-predicted prevalence, with 95% confidence intervals shown as error bars or shaded ribbon respectivly. For visualization of the *Season*× *Bodyweight* interaction, the range of standardized Body weight was restricted to the 2.5th and 97.5th percentiles of the observed data to avoid extrapolation at the tails. Significant pairwise differences are indicated by brackets and asterisks (* *p* < 0.05, ** *p* < 0.01, *** *p* < 0.001).

Sex was retained as a significant main predictor in the final GLM. Estimated marginal means indicated an adjusted predicted prevalence of 8.3% (95% *CI* : 4.55 −14.81) in females and 23.7% (95% *CI* : 17.01 −32.01) in males. The corresponding contrast showed that the odds of PUUV positivity were significantly lower in females than in males (*OR* = 0.29, 95% *CI* : 0.16 − 0.55, *z* = 3.81, *p* < 0.001) (Figure 2 (b)).

Model-based predicted prevalence increased from 8.6% (95% *CI* : 3.6−19.17) in 2020 and 12.8% (95% *CI* : 7.64 *™* 20.68) in 2021 to 25.6% (95% *CI* : 17.62 *™* 35.58) in the decline phase 2022. Exploratory pairwise contrasts showed no significant difference between 2020 and 2021, whereas 2020 had a significantly lower odds of PUUV positivity than 2022 (*OR* = 0.27, 95% *CI* : 0.10− 0.77, *z* =− 2.47, *p* < 0.05) and also significantly lower 2021 than in 2022 (*OR* = 0.43, 95% *CI* : 0.23 −0.80, *z* = 2.64, *p* < 0.01) (Figure 2 (c)).

There was a *Season*× *Bodyweight* interaction (*β* = 1.176, *SE* = 0.369, *p* < 0.01), with Body weight positively associated with PUUV positivity in autumn (*β* = 1.053, 95%*CI* : 0.523 − −1.582) but not in spring (Figure 2 (c)).

Out of the 83 PUUV RNA-positive samples, 68 bank voles yielded sequences spanning a 685-nt fragment of the N-coding region (positions 370–1055 corresponding to reference sequence of PUUV strain Puu/Vindeln/L20Cg/83, Accession number Z48586.1). From our dataset, we identified 48 sequence types and used these for the reconstruction of the phylogenetic tree (Supplementary Table 4). Sequence data are available from GenBank under accession numbers PZ464017 - PZ464063. All sequences from both sampling sites cluster within the North-Scandinavian PUUV lineage (N-SCA) (Figure 3). Pairwise nucleotide divergence among the obtained haplotypes ranged from 0.0–10.4% (mean: 4.3%), whereas amino acid divergence was substantially lower, ranging from 0.0–2.6% (mean: 0.5%). No distinct phylogenetic separation by sampling location, season, or year was observed, and haplotypes from both the coastal area and Vindeln were intermingled throughout the N-SCA clade.

**Figure 3:**
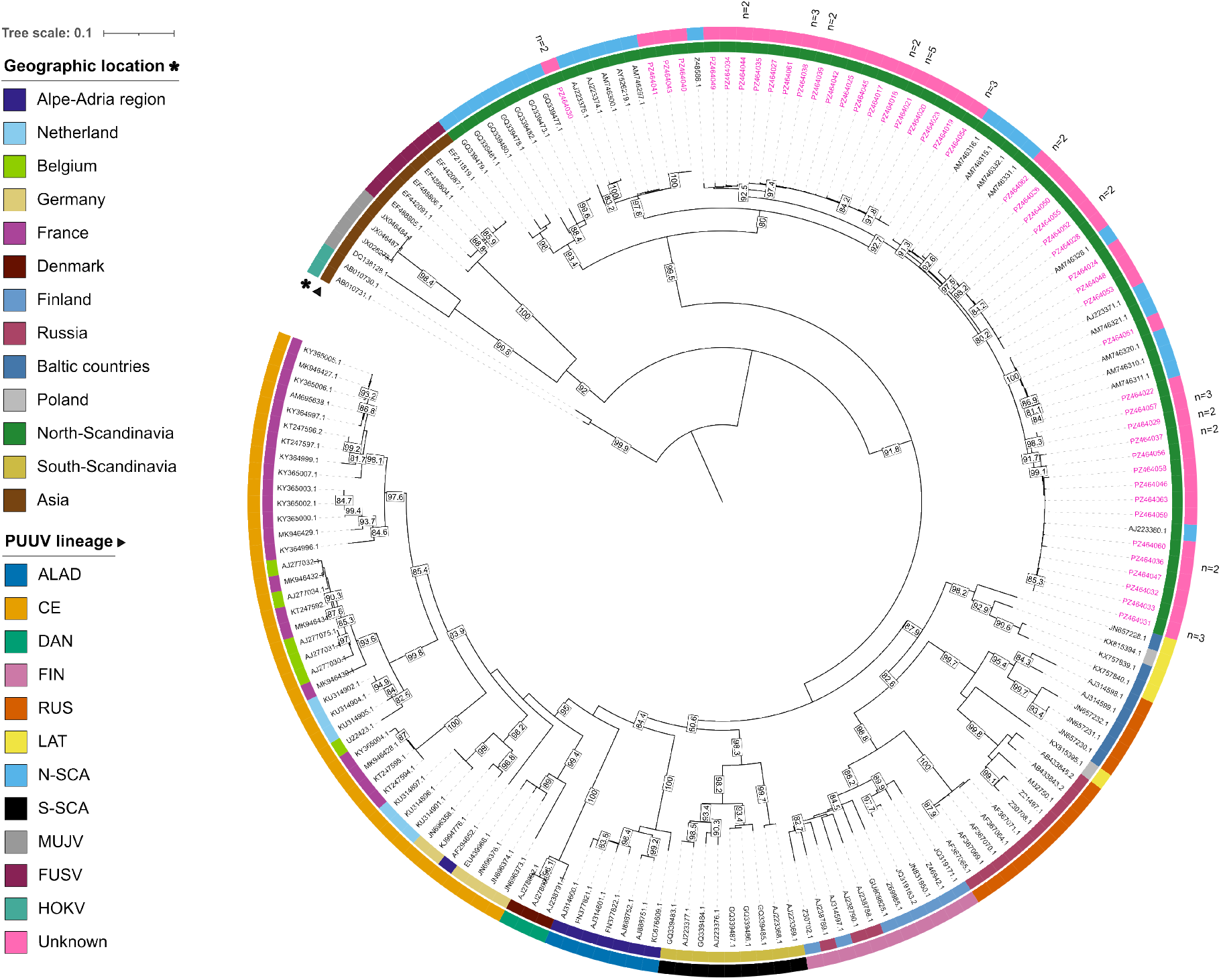
Maximum-likelihood tree based on parts (685nt) of the S segment specific N gene including bank voles >14.4g from Vindeln and coastal area from the years 2020-2022: The tree was constructed with IQ-TREE with the best-fit model (TIM2+F+I+G4) based on the Bayesian Information Criterion and 1,000 bootstrap replicates. We included 174 sequences, containing 286 parsimony-informative sites and 50 singletons. Only bootstrap values > 80% are shown. The tree was rooted using Hokkaido virus (HOKV) (GenBank AB010730.1 and AB010731.1). Sequences from this study are highlighted in pink as unknown lineage and cluster with the N-SCA PUUV lineage. Haplotype abundance (n).

Haversine distance calculations combined with Mantel testing demonstrated a significant positive correlation between genetic and geographic distances across all samples (Mantel *r* = 0.293, *p* ≤ 0.001).

### 3.3 Land-use

In the expert-based GLMMs, season was the only significant predictor of PUUV positivity at both spatial scales. PUUV positivity was higher in spring than in autumn at the 2.5× 2.5 *km*-scale (*β* = 1.662, SE = 0.340, *p* < 0.001, Table 3) and at the 200× 200 *m*-scale (*β* = 1.642, SE = 0.336, *p* < 0.001, Table 4). No clear associations between the evaluated land-use composition variables and PUUV positivity were identified at either spatial scale.

**Table 3.** Expert-based land-use GLMM for PUUV RNA positivity at the 2.5 × 2.5 *km* scale.

| Coefficient | Estimate | Std.Error | z value | Pr(> \|z\|) |
| --- | --- | --- | --- | --- |
| Intercept | -2.81 | 0.31 | -9.20 | <0.001 |
| SeasonSpring | 1.66 | 0.34 | 4.89 | <0.001 |
| Building | -0.28 | 0.34 | -0.82 | 0.411 |
| Clear-cut | 0.12 | 0.24 | 0.52 | 0.606 |
| Coniferous forest | -0.30 | 0.25 | -1.18 | 0.238 |
| Deciduous forest | 0.10 | 0.29 | 0.34 | 0.736 |
| Mixed forest | -0.45 | 0.27 | -1.68 | 0.093 |
| Wetland | 0.03 | 0.20 | 0.000 | 1.000 |
| Sampling intensity | 0.55 | 0.29 | 1.9 | 0.052 |

Estimates are on the logit scale. Continuous predictors were z-standardised prior to model fitting. Trapping plot was included as a random intercept Random intercept variance for trapping plot = 0.791, SD = 0.890. Number of observations = 519, number of trapping plots = 74.

**Table 4.**
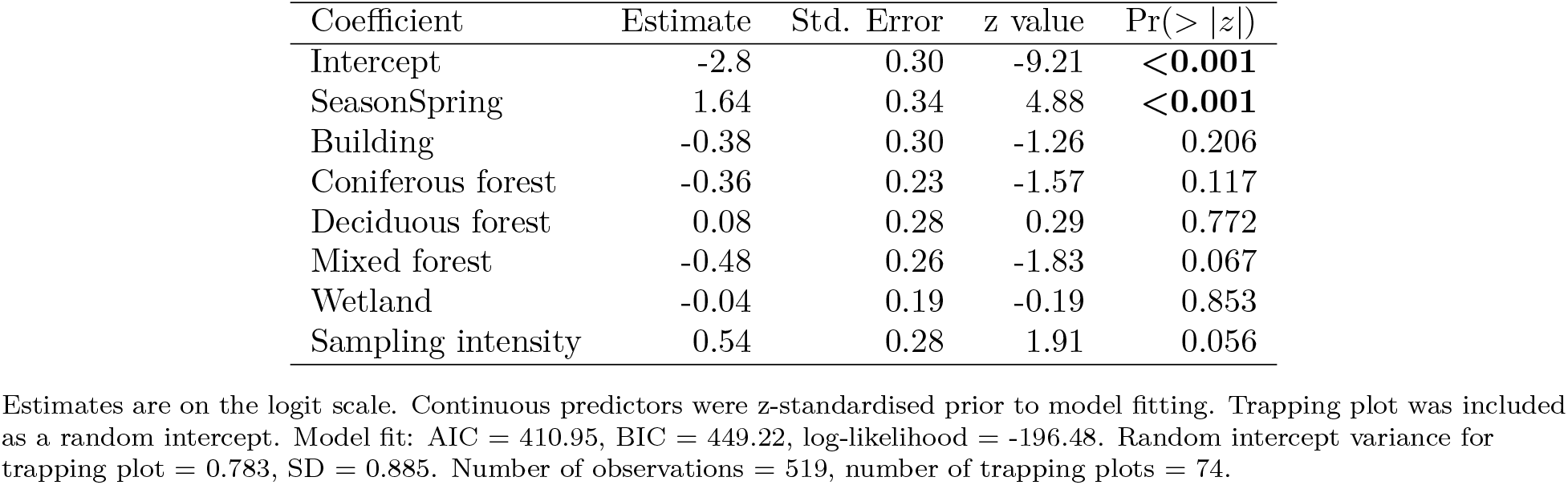
Expert-selected land-use GLMM for PUUV RNA positivity at the 200 × 200 *m* scale.

The explorative multi-scale landscape characterisation indicated substantial overlap in land-cover composition among PUUV-positive plots, PUUV-negative plots, and random background points (Supplementary Figure 2). Consistent with the GLMM results, compositional differences between groups were weak. No clear separation in land-cover structure was apparent relative to the background landscape, although PUUV-positive plots showed a slight tendency towards higher proportions of non-forest dry land-cover classes.

## 4 Discussion

This study showed, that PUUV occurrence in bank vole populations is mainly associated with seasonal and host-related factors. One of the main findings was the *Season* ×*Area* interaction, indicating that seasonal variation in PUUV positivity was not consistent across the two study areas despite rather close distance among these neighbouring areas (minimumm 3.7 km, maximum 109 km). PUUV occurrence was therefore not only related to season, but also to local population or environmental conditions that differed between areas.

Higher PUUV positivity in spring is consistent with previous studies reporting increased PUUV prevalence after winter (Khalil et al., 2019). Spring bank vole populations are largely composed of overwintered adults, which have had a longer cumulative opportunity for exposure to PUUV (Voutilainen et al., 2016). In contrast, autumn populations are typically dominated by juveniles and subadults born during the current breeding season, which are generally younger, lighter, and less likely to have encountered the virus (Olsson et al., 2002; Wang et al., 2023). This seasonal demographic turnover likely contributes to the lower apparent autumn prevalence, potentially reinforced by temporary maternal antibodies in juvenile bank voles (Kallio et al., 2006b). The significant *Season* ×*Bodyweight* interaction further supports this interpretation, indicating that the relationship between body weight and PUUV positivity differed between seasons. Rather than body weight acting as a direct causal factor, it likely reflects differences in age structure and accumulated exposure time, with heavier individuals in spring more likely representing overwintered adults. Previous work has also shown that PUUV infection may reduce overwinter survival (Kallio et al., 2007), which could further influence the composition of infected spring populations. The lower predicted PUUV positivity in spring in the Coastal area should however, be interpreted cautiously, as the limited spring sample size reduced precision and increased uncertainty around area-specific estimates.

PUUV positivity was also higher in males than in females, which alignes with previous serological stud-ies reporting male-biased hantavirus infection patterns across Europe (Bernshtein et al., 1999; Reil et al., 2017). This pattern is well-documented in Fennoscandian populations as well, where serological surveys have consistently shown higher antibody prevalence in males (Olsson et al., 2002; Voutilainen et al., 2012). This pattern may be explained by sex-specific behaviour, particularly larger home ranges (Escutenaire et al., 2002; Kozakiewicz et al., 2007), higher mobility and possible increased exposure probability (Korn, 1986) especially during the breeding season (Bernshtein et al., 1999). However, the observed sex effect should not be interpreted as an effect of sex alone. It probably reflects a combination of sex, age, body size, reproductive behaviour, movement, and social interactions.

Annual variation in PUUV positivity may be linked to the bank vole density over the course of the population cycle. PUUV occurrence increased over the study period and was significantly higher during the peak year 2021 compared to the beginning of the population cycle (increase year 2020). This is in line with the expectation that higher host density increases contact rates and environmental contamination, thereby facilitating virus transmission (Olsson et al., 2002). Previous serological studies have shown that PUUV infection dynamics are closely linked to cyclic fluctuations in bank vole density, with higher infection prevalence often occurring during high-density or peak phases of the host population cycle (Khalil et al., 2016; Voutilainen et al., 2016; Khalil et al., 2019). This is a pattern consistently observed also in Central Europe (Reil et al., 2017).

Phylogenetic analysis showed that all detected PUUV sequences clustered within the North-Scandinavian lineage, with clear separation from PUUV lineages reported from other geographic regions. This confirms that the viral population in the study area belongs to a well-established regional lineage (Castel et al., 2019), reinforcing previous findings from northern Sweden showing close phylogenetic relationships between PUUV sequences from human infections and wild-type bank vole isolates (Rosenbaum et al., 2024). Within the North-Scandinavian lineage, no distinct clustering by sampling area, season, or year was observed, indicating limited spatial and temporal structuring at the scale investigated. Instead, haplotypes were distributed across both study areas, suggesting shared transmission dynamics and host-mediated viral connectivity between the sampled bank vole populations. The substantially lower amino-acid divergence compared with nucleotide divergence suggests that the analysed N-coding fragment is highly conserved at the protein level, with many nucleotide substitutions likely being synonymous rather than resulting in amino-acid changes. In line with this, the significant correlation between genetic and geographic distance indicates isolation by distance, where viral genetic similarity decreases gradually with increasing spatial distance rather than forming discrete area-specific clusters. Overall, these findings point to relatively low phylogeographic structuring of PUUV within the study region. This is biologically plausible given the close proximity of the two sampling areas and the ecology of the reservoir host, the bank vole. Dispersal behaviour, particularly in males and subadult females seeking new territories or mating opportunities, can facilitate the movement of infected individuals across the landscape (Górska et al., 2025; Bernshtein et al., 1999). Such host-mediated connectivity may promote viral mixing and overlapping transmission networks, thereby preventing strong phylogeographic differentiation (Olsson et al., 2002).

Although local environmental conditions may shape PUUV transmission, as suggested by the *Season*×*Area* interaction, the expert-based land-use GLMMs did not identify significant associations between PUUV positivity and land-use composition at either the 2.5 ×2.5 km or 200× 200 m scale. Instead, season was the only significant predictor in these models, indicating that seasonal host–virus dynamics outweighed the explanatory contribution of the habitat variables included in this study. This suggests that temporal variation in host demography, accumulated exposure, and population turnover may be more influential for infection probability than broad-scale land-cover composition here.

In contrast to previous studies reporting associations between PUUV prevalence and habitat structure and quality (Khalil et al., 2016; Ecke et al., 2017; Khalil et al., 2019; Binder et al., 2020), no clear land-cover effects were detected here. This may partly reflect differences in land-cover representation between datasets, particularly regarding forest structure classes, such as clear-cuts, which may not be consistently resolved in the present data.

The explorative random-plot analysis showed broadly similar land-cover composition among PUUV-positive, PUUV-negative, and random background plots (Supplementary Figure 2). Overall, no clear compositional separation among groups was evident relative to the available landscape background. The non-forest dry land-cover class (NMD 118) was present at slightly greater proportions around PUUV-positive plots. However, this pattern was weak and should be interpreted only as descriptive ecological context, not as evidence of an independent land-cover effect. This is particularly relevant at the 200 ×200 m scale, where clear-cut, a related land-cover class, had to be excluded from the GLMM due to collinearity and elevated VIF values.

Taken together, our findings suggest that demography-based and seasonal factors as well as the phase of the vole cycle (Olsson et al., 2002; Voutilainen et al., 2016; Reil et al., 2017), were the primary drivers of PUUV positivity in the present study, while any potential land-cover signal was weak and not statistically resolved. More detailed spatial analysis incorporating specific land-cover classes, habitat configuration, and disturbance structure may provide further insight but were beyond the scope of this study, which focused primarily on host-related predictors of infection. Therefore, relevant ecological variation may occur at finer spatial scales or through variables not captured by land-cover classes, such as microhabitat structure, food availability, nesting sites, local climate conditions, or contact networks among bank voles (Khalil et al., 2017; Sipari et al., 2021; Olsson et al., 2005; Escutenaire et al., 2002).

Our findings are particularly relevant for PUUV spillover risk assessment, as they support previous studies identifying seasonal host–virus dynamics as a major component of infection risk in reservoir populations (Voutilainen et al., 2016; Wang et al., 2023; Khalil et al., 2019; Kallio et al., 2009). The higher PUUV positivity observed in spring suggests that the period following winter survival, but before substantial recruitment of uninfected juveniles, may represent an important window of increased virus circulation and potential human exposure in bank vole-suitable habitats, particularly during peak years of the bank vole population cycle as identified earlier (Khalil et al., 2019). Therefore, risk models based solely on static landscape composition may miss important temporal variation in reservoir infection dynamics.

## 5 Conclusions

This study shows that PUUV positivity in bank voles in northern Sweden is primarily shaped by seasonal host–virus dynamics, host demographic structure, and bank vole density as reflected in the phase of the vole cycle. Season was the most consistent predictor across analyses, with higher PUUV positivity in spring, suggesting that overwinter survival and accumulated exposure among bank voles play an important role in maintaining PUUV circulation in reservoir populations. In contrast, broad land-use composition at the analysed spatial scales showed no significant independent association with PUUV positivity, indicating that landscape effects may be weaker than seasonal dynamics or may operate through finer-scale ecological variables not captured by the available land-cover data.

Molecular phylogenetic analysis confirmed that the detected PUUV sequences belong to the North-Scandinavian lineage and showed limited spatial or temporal structuring within the study region. This suggests a connected regional viral population rather than clearly separated local viral foci. Overall, these findings highlight the importance of integrating seasonal reservoir dynamics, host population monitoring, landscape context, and viral genetic data in PUUV spillover risk assessment.

## Data Availability

All data produced in the present work are contained in the manuscript

## Acknowledgments

We thank Mikael Marberg (Swedish University of Agricultural Sciences (SLU)), Anna Johansson (SLU), and Sonya Juthberg (SLU) for their support with dissection work. Simone Lueert (Friedrich-Loeffler-Institut) is thanked for supporting the laboratory work.

## Financial disclosure

This research was supported by the European Union, grant agreement no. 101060568, project: BEPREP, and the Swedish Environmental Protection Agency through the Swedish Wildlife Management Fund (Grant No. 2020–00093).

## Conflict of interest

The authors declare no potential conflict of interest.

## Notes

### Competing Interest Statement

The authors have declared no competing interest.

