## Supplementary material for "Prevalence, Genetic Diversity, and Landscape Associations of *Orthohantavirus puumalaense* in Bank Voles (*Clethrionomys glareolus*) from Northern Sweden": Figure S1

### SUPPORTING INFORMATION

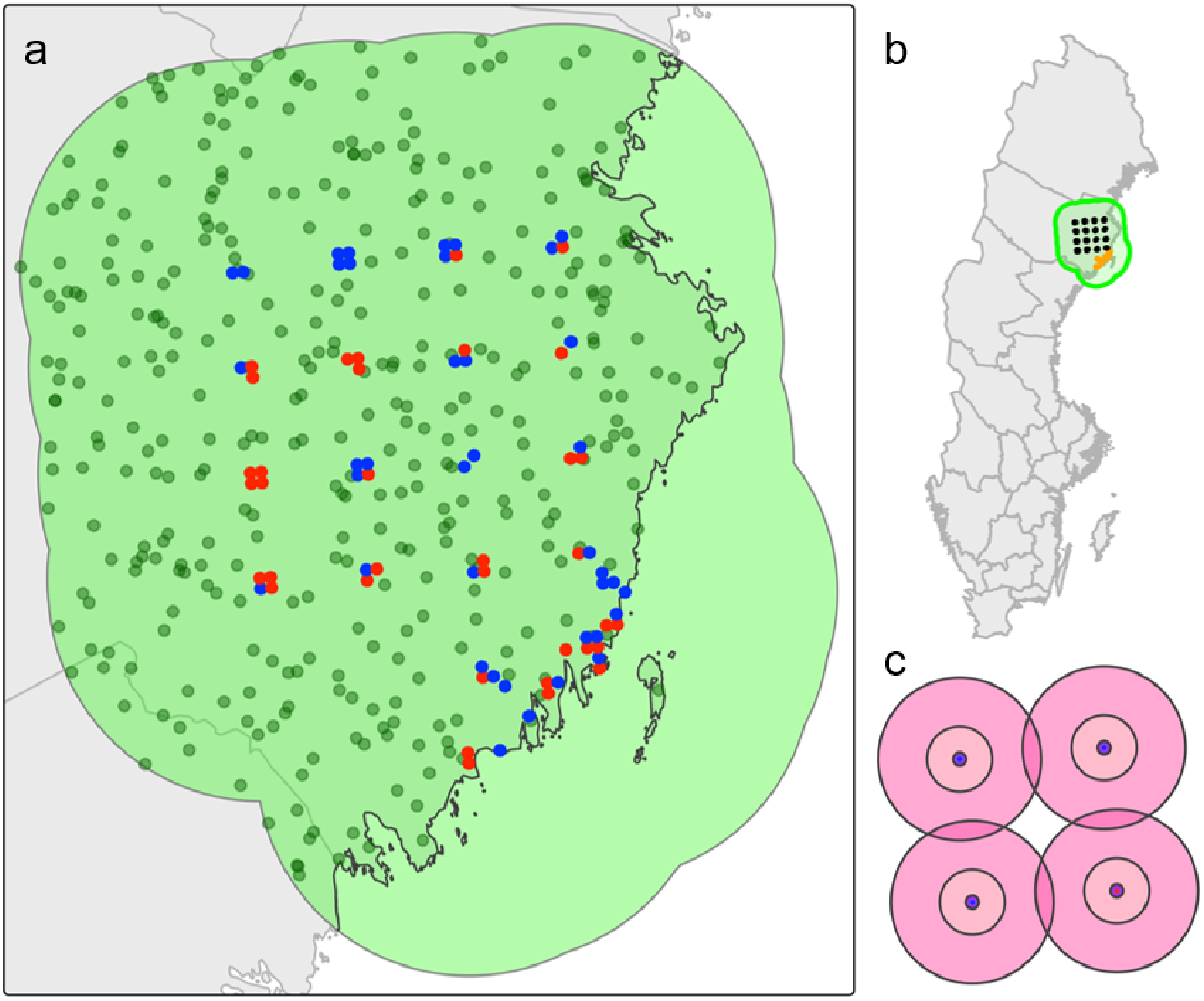


**FIGURE S1 Spatial context of Puumala virus (PUUV) sampling in northern Sweden.** A) Main study area map illustrating the spatial distribution of the bank vole trapping locations coloured by infection status (PUUV-positive = red, PUUV-negative = blue), alongside randomly seeded background points (dark green). B) Overview of the study area showing the location of the study area in northern Sweden, defined by a 50 km buffer around trapping plots in relation to the coastal and inland sampling regions (Vindeln = black and coastal area = orange). C) Example zoom-in of four selected trapping plots showing hierarchical landscape buffers used for land-cover composition analysis (113 m, 564 m, and 1410 m radii) around individual sampling locations.
