## Supplementary material for "Prevalence, Genetic Diversity, and Landscape Associations of *Orthohantavirus puumalaense* in Bank Voles (*Clethrionomys glareolus*) from Northern Sweden": Figure S2

### SUPPORTING INFORMATION

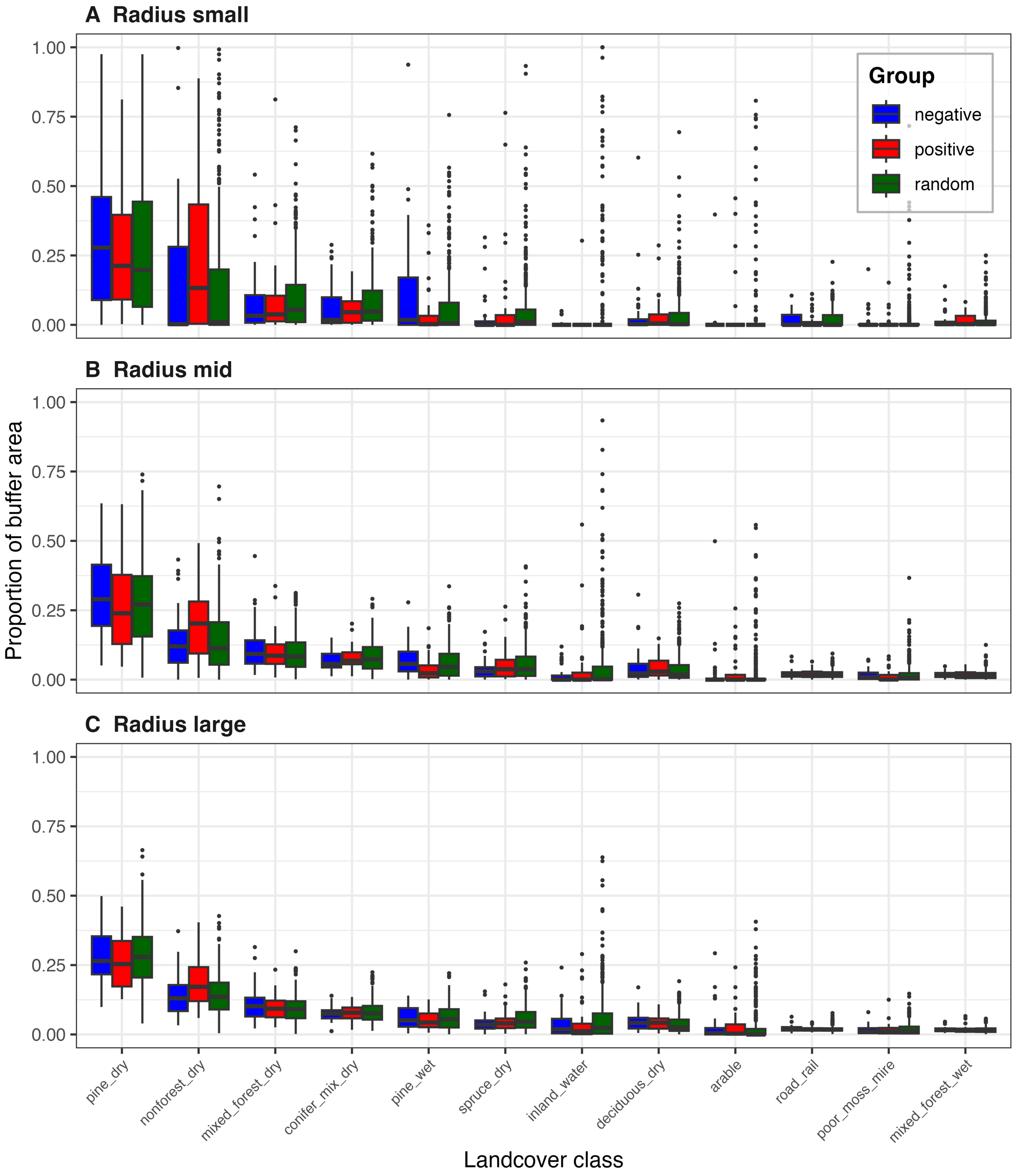


**FIGURE S2 Exploratory comparison of detailed landcover composition surrounding PUUV-positive bank vole plots, PUUV-negative plots, and randomly generated background plots across three spatial scales in northern Sweden.** Boxplots show the proportional cover of the twelve most abundant Swedish National Land Cover Database (NMD) classes within circular buffers around each plot. Random background plots were generated within the broader boreal study area to characterise underlying landscape composition independently of the spatial trapping design. (A) Small radius buffers (113 m). (B) Intermediate (mid) radius buffers (564 m). (C) Large radius buffers (1410 m). Red = PUUV-positive plots, blue = PUUV-negative plots, green = randomly seeded background plots. Results are descriptive and intended to visualise broad-scale landscape composition patterns across spatial scales
