## Supplementary material for "Prevalence, Genetic Diversity, and Landscape Associations of *Orthohantavirus puumalaense* in Bank Voles (*Clethrionomys glareolus*) from Northern Sweden": Table S1

### SUPPORTING INFORMATION

**TABLE** S1: Sample metadata for the 48 unique PUUV sequences included in the phylogenetic analysis.

| Sequence_ID | Geographic | Area | Year | Season | Sex | Age Category |
|---|---|---|---|---|---|---|
| KU0002995_Coast | North-Scandinavia | Coast | 2020 | Spring | Male | OWBR |
| KU0003148_Coast | North-Scandinavia | Coast | 2021 | Autumn | Male | ADUL |
| KU0003158_Coast | North-Scandinavia | Coast | 2021 | Autumn | Female | SUBA $\geq 14.4$ |
| KU0003159_Coast | North-Scandinavia | Coast | 2021 | Autumn | Male | SUBA $\geq 14.4$ |
| KU0003180_Coast | North-Scandinavia | Coast | 2021 | Autumn | Male | SUBA $\geq 14.4$ |
| KU0003194_Coast | North-Scandinavia | Coast | 2021 | Autumn | Male | ADUL |
| KU0003212_Coast | North-Scandinavia | Coast | 2021 | Autumn | Male | SUBA $\geq 14.4$ |
| KU0003634_Coast | North-Scandinavia | Coast | 2022 | Spring | Male | OWBR |
| KU0003685_Coast | North-Scandinavia | Coast | 2022 | Autumn | Female | SUBA $\geq 14.4$ |
| KU0003690_Coast | North-Scandinavia | Coast | 2022 | Autumn | Male | SUBA $\geq 14.4$ |
| VB0027796_Vindeln | North-Scandinavia | Vindeln | 2021 | Autumn | Male | ADUL |
| VB0027573_Vindeln | North-Scandinavia | Vindeln | 2021 | Spring | Male | OWBR |
| VB0027693_Vindeln | North-Scandinavia | Vindeln | 2021 | Autumn | Male | SUBA $\geq 14.4$ |
| VB0027602_Vindeln | North-Scandinavia | Vindeln | 2021 | Spring | Female | OWBR |
| VB0027630_Vindeln | North-Scandinavia | Vindeln | 2021 | Spring | Female | OWBR |
| VB0027631_Vindeln | North-Scandinavia | Vindeln | 2021 | Spring | Male | OWBR |
| VB0027636_Vindeln | North-Scandinavia | Vindeln | 2021 | Spring | Male | OWBR |
| VB0027647_Vindeln | North-Scandinavia | Vindeln | 2021 | Spring | Male | OWBR |
| VB0027648_Vindeln | North-Scandinavia | Vindeln | 2021 | Spring | Male | OWBR |
| VB0028884_Vindeln | North-Scandinavia | Vindeln | 2022 | Autumn | Female | SUBA $\geq 14.4$ |
| VB0028700_Vindeln | North-Scandinavia | Vindeln | 2022 | Spring | Male | OWBR |
| VB0028649_Vindeln | North-Scandinavia | Vindeln | 2022 | Spring | Male | OWBR |
| VB0028807_Vindeln | North-Scandinavia | Vindeln | 2022 | Spring | Male | OWBR |
| VB0028646_Vindeln | North-Scandinavia | Vindeln | 2022 | Spring | Male | OWBR |
| VB0028769_Vindeln | North-Scandinavia | Vindeln | 2022 | Spring | Male | OWBR |
| VB0028734_Vindeln | North-Scandinavia | Vindeln | 2022 | Spring | Male | OWBR |
| VB0028720_Vindeln | North-Scandinavia | Vindeln | 2022 | Spring | Female | OWBR |
| VB0028715_Vindeln | North-Scandinavia | Vindeln | 2022 | Spring | Male | OWBR |
| VB0028806_Vindeln | North-Scandinavia | Vindeln | 2022 | Spring | Male | OWBR |
| VB0028786_Vindeln | North-Scandinavia | Vindeln | 2022 | Spring | Male | OWBR |
| VB0028787_Vindeln | North-Scandinavia | Vindeln | 2022 | Spring | Male | OWBR |
| VB0029367_Vindeln | North-Scandinavia | Vindeln | 2022 | Autumn | Male | SUBA $\geq 14.4$ |
| VB0029393_Vindeln | North-Scandinavia | Vindeln | 2022 | Autumn | Female | SUBA $\geq 14.4$ |
| VB0028637_Vindeln | North-Scandinavia | Vindeln | 2022 | Spring | Male | OWBR |
| VB0028634_Vindeln | North-Scandinavia | Vindeln | 2022 | Spring | Female | OWBR |
| VB0028543_Vindeln | North-Scandinavia | Vindeln | 2021 | Autumn | Female | ADUL |
| VB0028500_Vindeln | North-Scandinavia | Vindeln | 2021 | Autumn | Male | SUBA $\geq 14.4$ |
| VB0028683_Vindeln | North-Scandinavia | Vindeln | 2022 | Spring | Male | OWBR |
| VB0028667_Vindeln | North-Scandinavia | Vindeln | 2022 | Spring | Male | OWBR |
| VB0028673_Vindeln | North-Scandinavia | Vindeln | 2022 | Spring | Male | OWBR |
| VB0028718_Vindeln | North-Scandinavia | Vindeln | 2022 | Spring | Female | OWBR |
| VB0028706_Vindeln | North-Scandinavia | Vindeln | 2022 | Spring | Male | OWBR |
| VB0028704_Vindeln | North-Scandinavia | Vindeln | 2022 | Spring | Male | OWBR |
| VB0028721_Vindeln | North-Scandinavia | Vindeln | 2022 | Spring | Male | OWBR |
| VB0028846_Vindeln | North-Scandinavia | Vindeln | 2022 | Spring | Male | OWBR |

| Sequence_ID | Geographic | Area | Year | Season | Sex | Age Category |
|---|---|---|---|---|---|---|
| VB0028809_Vindeln | North-Scandinavia | Vindeln | 2022 | Spring | Male | OWBR |
| VB0028811_Vindeln | North-Scandinavia | Vindeln | 2022 | Spring | Male | OWBR |
