## Supplementary material for "Prevalence, Genetic Diversity, and Landscape Associations of *Orthohantavirus puumalaense* in Bank Voles (*Clethrionomys glareolus*) from Northern Sweden": Table S2

### SUPPORTING INFORMATION

**TABLE** S2: NMD land-cover codes and corresponding Swedish and English class names.

| NMD_Code | Klass_swedish | clean_klass_swedish | Class_english |
|---|---|---|---|
| 3 | Åkermark | akermark | arable |
| 23 | Låg fjällskog på våtmark | lag_fjallskog_pa_vatmark | low_mountain_forest_wet |
| 43 | Låg fjällskog på fastmark | lag_fjallskog_pa_fastmark | low_mountain_forest_dry |
| 51 | Byggnad | byggnad | building |
| 52 | Anlagd mark, ej byggnad eller väg/järnväg | anlagd_mark_ej_byggnad_eller_vag_jarnvag | artificial |
| 53 | Väg eller järnväg | vag_eller_jarnvag | road_rail |
| 54 | Torvtäkt | torvtakt | peat_extraction |
| 61 | Inlandsvatten | inlandsvatten | inland_water |
| 62 | Hav | hav | sea |
| 111 | Tallskog på fastmark | tallskog_pa_fastmark | pine_dry |
| 112 | Granskog på fastmark | granskog_pa_fastmark | spruce_dry |
| 113 | Barrblandskog på fastmark | barrblandskog_pa_fastmark | conifer_mix_dry |
| 114 | Lövblandad barrskog på fastmark | lovblandad_barrskog_pa_fastmark | mixed_forest_dry |
| 115 | Triviallövskog på fastmark | triviallovskog_pa_fastmark | deciduous_dry |
| 116 | Ädellövskog på fastmark | adellovskog_pa_fastmark | broadleaf_dry |
| 117 | Triviallövskog med ädellövinslag på fastmark | triviallovskog_med_adellovinslag_pa_fastmark | mixed_deciduous_dry |
| 118 | Temporärt ej skog på fastmark | temporart_ej_skog_pa_fastmark | nonforest_dry |
| 121 | Tallskog på våtmark | tallskog_pa_vatmark | pine_wet |
| 122 | Granskog på våtmark | granskog_pa_vatmark | spruce_wet |
| 123 | Barrblandskog på våtmark | barrblandskog_pa_vatmark | conifer_mix_wet |
| 124 | Lövblandad barrskog på våtmark | lovblandad_barrskog_pa_vatmark | mixed_forest_wet |
| 125 | Triviallövskog på våtmark | triviallovskog_pa_vatmark | deciduous_wet |
| 126 | Ädellövskog på våtmark | adellovskog_pa_vatmark | broadleaf_wet |
| 127 | Triviallövskog med ädellövinslag på våtmark | triviallovskog_med_adellovinslag_pa_vatmark | mixed_deciduous_wet |
| 128 | Temporärt ej skog på våtmark | temporart_ej_skog_pa_vatmark | nonforest_wet |
| 200 | Öppen våtmark (underindelning saknas) | oppen_vatmark_underindelning_saknas | wet_open |
| 211 | Buskmyr | buskmyr | shrub_mire |
| 212 | Ristuvemyr | ristuvemyr | brush_mire |
| 213 | Fastmattemyr, mager | fastmattemyr_mager | poor_moss_mire |
| 214 | Fastmattemyr, frodig | fastmattemyr_frodig | lush_moss_mire |
| 215 | Sumpkärr | sumpkarr | swamp |
| 216 | Mjukmattemyr | mjukmattemyr | soft_moss_mire |
| 217 | Lösbottenmyr | losbottenmyr | loose_mire |
| 218 | Övrig öppen myr | ovrig_oppen_myr | other_mire |
| 221 | Våtmark med buskar | vatmark_med_buskar | wet_shrub |
| 222 | Risdominerad våtmark | risdominerad_vatmark | bush_wet |
| 223 | Gräsdominerad våtmark, mager | grasdominerad_vatmark_mager | poor_grass_wet |
| 224 | Gräsdominerad våtmark, frodvuxen | grasdominerad_vatmark_frodvuxen | lush_grass_wet |

Continued on next page

**TABLE** S2: NMD land-cover codes and corresponding Swedish and English class names (continued).

| NMD_Code | Klass_swedish | clean_klass_swedish | Class_english |
|---|---|---|---|
| 225 | Gräsdominerad våtmark, högvuxen | grasdominerad_vatmark_hogvuxen | tall_grass_wet |
| 226 | Mossdominerad våtmark | mossdominerad_vatmark | moss_wet |
| 227 | Våtmark utan växttäcke | vatmark_utan_vaxttacke | bare_wetland |
| 228 | Övrig öppen våtmark | ovrig_oppen_vatmark | other_wet_open |
| 411 | Öppen fastmark utan vegetation (ej glaciär eller varaktigt snöfält) | oppen_fastmark_utan_vegetation ej_glaciar_eller_varaktigt_snofalt | bare_rock |
| 4211 | Torr buskdominerad mark | torr_buskdominerad_mark | dry_shrub |
| 4212 | Frisk buskdominerad mark | frisk_buskdominerad_mark | fresh_shrub |
| 4213 | Frisk-fuktig buskdominerad mark | frisk_fuktig_buskdominerad_mark | moist_shrub |
| 4221 | Torr risdominerad mark | torr_risdominerad_mark | dry_bush |
| 4222 | Frisk risdominerad mark | frisk_risdominerad_mark | fresh_bush |
| 4223 | Frisk-fuktig risdominerad mark | frisk_fuktig_risdominerad_mark | moist_bush |
| 4231 | Torr gräsdominerad mark | torr_grasdominerad_mark | dry_grass |
| 4232 | Frisk gräsdominerad mark | frisk_grasdominerad_mark | fresh_grass |
| 4233 | Frisk-fuktig gräsdominerad mark | frisk_fuktig_grasdominerad_mark | moist_grass |
