## Supplementary material for "Prevalence, Genetic Diversity, and Landscape Associations of *Orthohantavirus puumalaense* in Bank Voles (*Clethrionomys glareolus*) from Northern Sweden": Table S3

### SUPPORTING INFORMATION

**TABLE** S3: Summary of PUUV samples by trapping plot.

| Area | TOPO | Lat | Lon | 2020 | 2021 | 2022 | 2023 | Spring | Autumn | Males | Females | Juveniles | Adults | Mean BW | PUUV+ | PUUV- | Total |
|---|---|---|---|---|---|---|---|---|---|---|---|---|---|---|---|---|---|
| Coast | 20K3B1212 | 63.644 | 19.968 | 4 | 1 | 8 | 0 | 1 | 12 | 6 | 7 | 3 | 10 | 162.9 | 1 | 12 | 13 |
| Coast | 20K3B3712 | 63.666 | 19.972 | 0 | 6 | 6 | 0 | 0 | 12 | 8 | 4 | 0 | 12 | 177.2 | 1 | 11 | 12 |
| Coast | 20K3C3737 | 63.661 | 20.123 | 1 | 3 | 1 | 0 | 0 | 5 | 2 | 3 | 1 | 4 | 156.8 | 0 | 5 | 5 |
| Coast | 20K5E1212 | 63.724 | 20.284 | 0 | 4 | 2 | 0 | 0 | 6 | 4 | 2 | 2 | 4 | 149.8 | 0 | 6 | 6 |
| Coast | 20K6D3712 | 63.794 | 20.194 | 0 | 3 | 0 | 0 | 0 | 3 | 0 | 3 | 2 | 1 | 148.0 | 0 | 3 | 3 |
| Coast | 20K6F1212 | 63.765 | 20.392 | 5 | 3 | 9 | 0 | 3 | 14 | 10 | 7 | 3 | 14 | 182.6 | 4 | 13 | 17 |
| Coast | 20K6F3712 | 63.788 | 20.396 | 4 | 5 | 5 | 0 | 2 | 12 | 7 | 7 | 2 | 12 | 175.6 | 1 | 13 | 14 |
| Coast | 20K6F3737 | 63.786 | 20.446 | 2 | 0 | 2 | 0 | 2 | 2 | 2 | 2 | 0 | 4 | 220.2 | 0 | 4 | 4 |
| Coast | 20K7C1212 | 63.819 | 20.096 | 12 | 24 | 3 | 0 | 5 | 34 | 23 | 16 | 5 | 34 | 188.7 | 12 | 27 | 39 |
| Coast | 20K7C1237 | 63.818 | 20.147 | 5 | 0 | 6 | 0 | 2 | 9 | 4 | 7 | 1 | 10 | 174.6 | 0 | 11 | 11 |
| Coast | 20K7C3712 | 63.842 | 20.099 | 1 | 1 | 0 | 0 | 0 | 2 | 0 | 2 | 0 | 2 | 169.5 | 0 | 2 | 2 |
| Coast | 20K7H1237 | 63.802 | 20.652 | 2 | 0 | 2 | 0 | 2 | 2 | 4 | 0 | 0 | 4 | 224.2 | 1 | 3 | 4 |
| Coast | 20K7H3737 | 63.824 | 20.656 | 2 | 0 | 2 | 0 | 2 | 2 | 2 | 2 | 0 | 4 | 241.5 | 0 | 4 | 4 |
| Coast | 20K8G1212 | 63.852 | 20.508 | 1 | 0 | 6 | 0 | 6 | 1 | 3 | 4 | 0 | 7 | 249.4 | 1 | 6 | 7 |
| Coast | 20K8H1212 | 63.848 | 20.610 | 0 | 0 | 6 | 0 | 6 | 0 | 4 | 2 | 0 | 6 | 240.5 | 1 | 5 | 6 |
| Coast | 20K8H1237 | 63.847 | 20.660 | 1 | 0 | 8 | 0 | 8 | 1 | 6 | 3 | 0 | 9 | 248.2 | 1 | 8 | 9 |
| Coast | 20K8H3712 | 63.871 | 20.613 | 2 | 0 | 0 | 0 | 0 | 2 | 1 | 1 | 0 | 2 | 213.5 | 0 | 2 | 2 |
| Coast | 20K8H3737 | 63.869 | 20.664 | 1 | 0 | 4 | 0 | 4 | 1 | 3 | 2 | 1 | 4 | 211.0 | 0 | 5 | 5 |
| Coast | 20K9I1212 | 63.890 | 20.719 | 3 | 0 | 0 | 0 | 0 | 3 | 3 | 0 | 2 | 1 | 149.7 | 1 | 2 | 3 |
| Coast | 20K9I1237 | 63.888 | 20.769 | 0 | 0 | 2 | 0 | 2 | 0 | 2 | 0 | 0 | 2 | 234.5 | 2 | 0 | 2 |
| Coast | 20K9I3737 | 63.910 | 20.773 | 0 | 0 | 3 | 0 | 3 | 0 | 2 | 1 | 0 | 3 | 224.3 | 0 | 3 | 3 |
| Coast | 21K0J3712 | 63.953 | 20.832 | 0 | 1 | 2 | 0 | 3 | 0 | 2 | 1 | 0 | 3 | 251.7 | 0 | 3 | 3 |
| Coast | 21K1I1212 | 63.979 | 20.734 | 1 | 1 | 0 | 0 | 2 | 0 | 2 | 0 | 0 | 2 | 213.5 | 0 | 2 | 2 |
| Coast | 21K1I1237 | 63.977 | 20.785 | 0 | 0 | 2 | 0 | 2 | 0 | 1 | 1 | 0 | 2 | 213.5 | 0 | 2 | 2 |
| Coast | 21K1I3712 | 64.001 | 20.738 | 0 | 2 | 0 | 0 | 2 | 0 | 1 | 1 | 0 | 2 | 308.0 | 0 | 2 | 2 |
| Vindeln | 21J2C1212 | 64.070 | 19.109 | 0 | 8 | 3 | 1 | 3 | 9 | 8 | 4 | 3 | 9 | 179.3 | 0 | 12 | 12 |
| Vindeln | 21J2C1237 | 64.069 | 19.160 | 1 | 4 | 0 | 1 | 0 | 6 | 6 | 0 | 3 | 3 | 162.5 | 1 | 5 | 6 |
| Vindeln | 21J2C3712 | 64.092 | 19.111 | 0 | 2 | 8 | 0 | 6 | 4 | 8 | 2 | 0 | 10 | 214.1 | 3 | 7 | 10 |
| Vindeln | 21J2C3737 | 64.091 | 19.163 | 0 | 4 | 3 | 0 | 3 | 4 | 5 | 2 | 0 | 7 | 205.6 | 1 | 6 | 7 |
| Vindeln | 21J2H1212 | 64.057 | 19.620 | 0 | 0 | 2 | 0 | 2 | 0 | 2 | 0 | 0 | 2 | 276.5 | 1 | 1 | 2 |
| Vindeln | 21J2H3712 | 64.080 | 19.623 | 2 | 0 | 2 | 0 | 2 | 2 | 2 | 2 | 1 | 3 | 173.0 | 0 | 4 | 4 |
| Vindeln | 21J2H3737 | 64.078 | 19.674 | 2 | 2 | 2 | 0 | 4 | 2 | 5 | 1 | 0 | 6 | 235.5 | 3 | 3 | 6 |
| Vindeln | 21J7C1212 | 64.294 | 19.135 | 0 | 2 | 14 | 0 | 4 | 12 | 11 | 5 | 0 | 16 | 187.4 | 4 | 12 | 16 |
| Vindeln | 21J7C1237 | 64.293 | 19.187 | 2 | 4 | 4 | 0 | 2 | 8 | 6 | 4 | 1 | 9 | 184.4 | 1 | 9 | 10 |
| Vindeln | 21J7C3712 | 64.316 | 19.138 | 2 | 3 | 10 | 0 | 4 | 11 | 11 | 4 | 1 | 14 | 181.7 | 2 | 13 | 15 |
| Vindeln | 21J7C3737 | 64.315 | 19.190 | 0 | 8 | 4 | 0 | 0 | 12 | 8 | 4 | 5 | 7 | 139.2 | 1 | 11 | 12 |
| Vindeln | 21J7H1212 | 64.281 | 19.651 | 0 | 2 | 1 | 0 | 3 | 0 | 3 | 0 | 0 | 3 | 242.0 | 0 | 3 | 3 |
| Vindeln | 21J7H1237 | 64.280 | 19.702 | 1 | 4 | 3 | 0 | 7 | 1 | 4 | 4 | 0 | 8 | 232.6 | 1 | 7 | 8 |
| Vindeln | 21J7H3712 | 64.304 | 19.654 | 1 | 0 | 1 | 0 | 1 | 1 | 2 | 0 | 1 | 1 | 165.5 | 0 | 2 | 2 |
| Vindeln | 21J7H3737 | 64.302 | 19.705 | 0 | 0 | 1 | 0 | 1 | 0 | 1 | 0 | 0 | 1 | 255.0 | 0 | 1 | 1 |
| Vindeln | 21K2C1212 | 64.043 | 20.130 | 0 | 0 | 4 | 0 | 4 | 0 | 4 | 0 | 0 | 4 | 262.8 | 0 | 4 | 4 |
| Vindeln | 21K2C1237 | 64.042 | 20.181 | 0 | 1 | 6 | 0 | 7 | 0 | 3 | 4 | 0 | 7 | 234.6 | 5 | 2 | 7 |
| Vindeln | 21K2C3737 | 64.064 | 20.185 | 1 | 1 | 2 | 0 | 3 | 1 | 2 | 2 | 1 | 3 | 213.0 | 2 | 2 | 4 |
| Vindeln | 21K2H3712 | 64.049 | 20.644 | 1 | 0 | 2 | 0 | 2 | 1 | 2 | 1 | 1 | 2 | 216.0 | 1 | 2 | 3 |
| Vindeln | 21K2H3737 | 64.048 | 20.695 | 4 | 3 | 0 | 1 | 3 | 5 | 7 | 1 | 0 | 8 | 208.6 | 0 | 8 | 8 |
| Vindeln | 21K7C1212 | 64.267 | 20.165 | 0 | 0 | 3 | 0 | 3 | 0 | 0 | 3 | 0 | 3 | 239.0 | 0 | 3 | 3 |
| Vindeln | 21K7C3737 | 64.288 | 20.220 | 1 | 0 | 0 | 0 | 0 | 1 | 1 | 0 | 0 | 1 | 150.0 | 0 | 1 | 1 |
| Vindeln | 21K7H1212 | 64.250 | 20.679 | 0 | 0 | 1 | 0 | 1 | 0 | 1 | 0 | 0 | 1 | 217.0 | 1 | 0 | 1 |

Continued on next page

**TABLE** S3: Summary of PUUV samples by trapping plot (continued).

| Area | TOPO | Lat | Lon | 2020 | 2021 | 2022 | 2023 | Spring | Autumn | Males | Females | Juveniles | Adults | Mean BW | PUUV+ | PUUV- | Total |
|---|---|---|---|---|---|---|---|---|---|---|---|---|---|---|---|---|---|
| Vindeln | 21K7H1237 | 64.249 | 20.731 | 1 | 5 | 6 | 0 | 11 | 1 | 7 | 5 | 0 | 12 | 259.2 | 5 | 7 | 12 |
| Vindeln | 21K7H3737 | 64.271 | 20.735 | 5 | 3 | 2 | 0 | 5 | 5 | 5 | 5 | 2 | 8 | 198.0 | 0 | 10 | 10 |
| Vindeln | 22J2C1237 | 64.516 | 19.215 | 1 | 1 | 2 | 0 | 3 | 1 | 1 | 3 | 0 | 4 | 237.8 | 2 | 2 | 4 |
| Vindeln | 22J2C3712 | 64.540 | 19.166 | 1 | 0 | 0 | 0 | 0 | 1 | 0 | 1 | 0 | 1 | 219.0 | 0 | 1 | 1 |
| Vindeln | 22J2C3737 | 64.539 | 19.218 | 1 | 4 | 2 | 0 | 6 | 1 | 5 | 2 | 1 | 6 | 219.7 | 5 | 2 | 7 |
| Vindeln | 22J2H1237 | 64.504 | 19.734 | 0 | 1 | 1 | 1 | 3 | 0 | 3 | 0 | 0 | 3 | 226.0 | 1 | 2 | 3 |
| Vindeln | 22J2H3712 | 64.527 | 19.685 | 0 | 1 | 1 | 1 | 3 | 0 | 1 | 2 | 0 | 3 | 234.3 | 1 | 2 | 3 |
| Vindeln | 22J2H3737 | 64.526 | 19.737 | 2 | 7 | 1 | 0 | 8 | 2 | 7 | 3 | 0 | 10 | 252.6 | 7 | 3 | 10 |
| Vindeln | 22J7C1212 | 64.742 | 19.191 | 1 | 0 | 0 | 0 | 0 | 1 | 0 | 1 | 0 | 1 | 156.0 | 0 | 1 | 1 |
| Vindeln | 22J7C1237 | 64.740 | 19.243 | 0 | 1 | 0 | 0 | 1 | 0 | 0 | 1 | 1 | 0 | 119.0 | 0 | 1 | 1 |
| Vindeln | 22J7H1212 | 64.729 | 19.714 | 0 | 0 | 1 | 0 | 1 | 0 | 1 | 0 | 0 | 1 | 236.0 | 0 | 1 | 1 |
| Vindeln | 22J7H1237 | 64.727 | 19.767 | 2 | 0 | 1 | 0 | 1 | 2 | 2 | 1 | 0 | 3 | 226.3 | 0 | 3 | 3 |
| Vindeln | 22J7H3712 | 64.751 | 19.717 | 0 | 1 | 1 | 0 | 2 | 0 | 1 | 1 | 0 | 2 | 271.0 | 0 | 2 | 2 |
| Vindeln | 22J7H3737 | 64.750 | 19.770 | 1 | 0 | 3 | 1 | 4 | 1 | 3 | 2 | 1 | 4 | 171.8 | 0 | 5 | 5 |
| Vindeln | 22K2C1212 | 64.490 | 20.201 | 1 | 0 | 0 | 0 | 0 | 1 | 1 | 0 | 0 | 1 | 162.0 | 0 | 1 | 1 |
| Vindeln | 22K2C1237 | 64.489 | 20.253 | 2 | 0 | 2 | 0 | 3 | 1 | 2 | 2 | 0 | 4 | 244.8 | 0 | 4 | 4 |
| Vindeln | 22K2C3737 | 64.511 | 20.256 | 1 | 0 | 4 | 0 | 4 | 1 | 2 | 3 | 1 | 4 | 179.6 | 1 | 4 | 5 |
| Vindeln | 22K2H1212 | 64.474 | 20.719 | 5 | 0 | 1 | 0 | 1 | 5 | 3 | 3 | 3 | 3 | 140.2 | 1 | 5 | 6 |
| Vindeln | 22K2H3737 | 64.494 | 20.775 | 4 | 2 | 0 | 0 | 2 | 4 | 2 | 4 | 0 | 6 | 189.8 | 0 | 6 | 6 |
| Vindeln | 22K7C1212 | 64.714 | 20.237 | 1 | 5 | 3 | 0 | 2 | 7 | 5 | 4 | 1 | 8 | 187.8 | 0 | 9 | 9 |
| Vindeln | 22K7C1237 | 64.712 | 20.290 | 0 | 6 | 11 | 0 | 0 | 17 | 7 | 10 | 0 | 17 | 172.8 | 3 | 14 | 17 |
| Vindeln | 22K7C3712 | 64.736 | 20.241 | 0 | 15 | 9 | 0 | 2 | 22 | 10 | 14 | 9 | 15 | 164.9 | 0 | 24 | 24 |
| Vindeln | 22K7C3737 | 64.735 | 20.293 | 0 | 1 | 0 | 0 | 0 | 1 | 0 | 1 | 0 | 1 | 164.0 | 0 | 1 | 1 |
| Vindeln | 22K7H1212 | 64.697 | 20.760 | 2 | 9 | 2 | 0 | 0 | 13 | 8 | 5 | 2 | 11 | 168.8 | 0 | 13 | 13 |
| Vindeln | 22K7H1237 | 64.696 | 20.812 | 4 | 18 | 14 | 4 | 8 | 32 | 24 | 16 | 5 | 35 | 178.8 | 4 | 36 | 40 |
| Vindeln | 22K7H3737 | 64.718 | 20.816 | 0 | 4 | 1 | 0 | 1 | 4 | 4 | 1 | 1 | 4 | 189.2 | 0 | 5 | 5 |
