## Supplementary material for "Prevalence, Genetic Diversity, and Landscape Associations of *Orthohantavirus puumalaense* in Bank Voles (*Clethrionomys glareolus*) from Northern Sweden": Table S4

### SUPPORTING INFORMATION

**TABLE** S4: Sequences included in phylogenetic analysis: Accession numbers with geographic origin and PUUV lineage information.

| Accession | Geographic | Lineage |
|---|---|---|
| AJ888752.1 | Alpe-Adria region | ALAD |
| AJ888751.1 | Alpe-Adria region | ALAD |
| KC676609.1 | Alpe-Adria region | ALAD |
| AJ314600.1 | Alpe-Adria region | ALAD |
| AJ314601.1 | Alpe-Adria region | ALAD |
| FN377822.1 | Alpe-Adria region | ALAD |
| FN377821.1 | Alpe-Adria region | ALAD |
| AF294652.1 | Alpe-Adria region | CE |
| KU314901.1 | Netherland | CE |
| KU314897.1 | Netherland | CE |
| KU314896.1 | Netherland | CE |
| KU314905.1 | Netherland | CE |
| KU314902.1 | Netherland | CE |
| KU314904.1 | Netherland | CE |
| U22423.1 | Belgium | CE |
| AJ277075.1 | Belgium | CE |
| AJ277034.1 | Belgium | CE |
| AJ277032.1 | Belgium | CE |
| AJ277031.1 | Belgium | CE |
| AJ277030.1 | Belgium | CE |
| EU439968.1 | Germany | CE |
| JN696358.1 | Germany | CE |
| JN696373.1 | Germany | CE |
| JN696374.1 | Germany | CE |
| JN696376.1 | Germany | CE |
| KJ994776.1 | Germany | CE |
| AM695638.1 | France | CE |
| KT247592.1 | France | CE |
| KT247596.2 | France | CE |
| KT247597.1 | France | CE |
| KT247594.1 | France | CE |
| KT247595.1 | France | CE |
| KY364996.1 | France | CE |
| KY365003.1 | France | CE |
| KY365002.1 | France | CE |
| KY365000.1 | France | CE |
| KY365007.1 | France | CE |
| KY364997.1 | France | CE |
| KY364999.1 | France | CE |
| KY365004.1 | France | CE |
| MK946428.1 | France | CE |
| KY365005.1 | France | CE |
| KY365006.1 | France | CE |
| MK946427.1 | France | CE |
| MK946429.1 | France | CE |

Continued on next page

**Table S4 continued from previous page**

| Accession | Geographic | Lineage |
|---|---|---|
| MK946434.1 | France | CE |
| MK946432.1 | France | CE |
| MK946430.1 | France | CE |
| AJ238791.1 | Denmark | DAN |
| AJ278092.1 | Denmark | DAN |
| AJ278093.1 | Denmark | DAN |
| Z30702.1 | Finland | FIN |
| GU808825.1 | Finland | FIN |
| AJ314597.1 | Finland | FIN |
| Z46942.1 | Finland | FIN |
| Z69985.1 | Finland | FIN |
| JQ319171.1 | Finland | FIN |
| JQ319163.2 | Finland | FIN |
| JN831950.1 | Finland | FIN |
| AJ238790.1 | Russia | FIN |
| AJ238788.1 | Russia | FIN |
| AJ238789.1 | Russia | FIN |
| AF367064.1 | Russia | RUS |
| AF367065.1 | Russia | RUS |
| AF367069.1 | Russia | RUS |
| AF367070.1 | Russia | RUS |
| AF367071.1 | Russia | RUS |
| M32750.1 | Russia | RUS |
| AB433843.2 | Russia | RUS |
| AB433845.2 | Russia | RUS |
| Z30708.1 | Russia | RUS |
| Z21497.1 | Russia | RUS |
| AJ314599.1 | Baltic countries | RUS |
| AJ314598.1 | Baltic countries | RUS |
| JN657230.1 | Latvia | RUS |
| JN657231.1 | Latvia | RUS |
| JN657232.1 | Latvia | RUS |
| JN657228.1 | Latvia | LAT |
| KX757840.1 | Lithuania | LAT |
| KX757839.1 | Lithuania | LAT |
| KX815394.1 | Poland | LAT |
| KX815395.1 | Poland | LAT |
| GQ339477.1 | North-Scandinavia | N-SCA |
| AM746297.1 | North-Scandinavia | N-SCA |
| AM746300.1 | North-Scandinavia | N-SCA |
| AM746310.1 | North-Scandinavia | N-SCA |
| AM746311.1 | North-Scandinavia | N-SCA |
| AM746315.1 | North-Scandinavia | N-SCA |
| AM746316.1 | North-Scandinavia | N-SCA |
| GQ339480.1 | North-Scandinavia | N-SCA |
| GQ339478.1 | North-Scandinavia | N-SCA |
| GQ339482.1 | North-Scandinavia | N-SCA |

Continued on next page

**Table S4 continued from previous page**

| Accession | Geographic | Lineage |
|---|---|---|
| GQ339473.1 | North-Scandinavia | N-SCA |
| GQ339481.1 | North-Scandinavia | N-SCA |
| GQ339479.1 | North-Scandinavia | N-SCA |
| AM746320.1 | North-Scandinavia | N-SCA |
| AM746321.1 | North-Scandinavia | N-SCA |
| AM746328.1 | North-Scandinavia | N-SCA |
| AJ223371.1 | North-Scandinavia | N-SCA |
| AJ223374.1 | North-Scandinavia | N-SCA |
| AJ223375.1 | North-Scandinavia | N-SCA |
| AJ223380.1 | North-Scandinavia | N-SCA |
| Z48586.1 | North-Scandinavia | N-SCA |
| AM746331.1 | North-Scandinavia | N-SCA |
| AM746332.1 | North-Scandinavia | N-SCA |
| AY526219.1 | North-Scandinavia | N-SCA |
| GQ339483.1 | South-Scandinavia | S-SCA |
| GQ339484.1 | South-Scandinavia | S-SCA |
| GQ339485.1 | South-Scandinavia | S-SCA |
| GQ339487.1 | South-Scandinavia | S-SCA |
| GQ339486.1 | South-Scandinavia | S-SCA |
| AJ223368.1 | South-Scandinavia | S-SCA |
| AJ223369.1 | South-Scandinavia | S-SCA |
| AJ223376.1 | South-Scandinavia | S-SCA |
| AJ223377.1 | South-Scandinavia | S-SCA |
| DQ138128.1 | Asia | MUJV |
| JX028273.1 | Asia | MUJV |
| JX046484.1 | Asia | MUJV |
| JX046487.1 | Asia | MUJV |
| EF488805.1 | Asia | FUSV |
| EF211819.1 | Asia | FUSV |
| EF488804.1 | Asia | FUSV |
| EF488806.1 | Asia | FUSV |
| EF442087.1 | Asia | FUSV |
| EF442091.1 | Asia | FUSV |
| AB010730.1 | Asia | HOKV |
| AB010731.1 | Asia | HOKV |
